# Chromosomal copy number variations in miscarriages and the geneic counseling of recurrent pregnancy loss

**DOI:** 10.1101/2023.10.06.23292110

**Authors:** Hui Hu, Jin Li, Jie Duan, Li Yu, Guangming Ye, Na Gao, Na Yang, Xueping Qiu, Xin Jin, Shuyang Sheng, Yating Cheng, Yuanzhen Zhang, Jianhong Ma, Fang Zheng

## Abstract

The purpose of this study was to explore the copy number variations (CNVs) associated with miscarriage. A total of 662 specimens of aborted embryonic tissue and 54 samples from peripheral blood were collected. Next generation sequencing for CNV analysis was performed to determine the type and clinical significance of possible CNVs, and relevant medical records were collected. Autosomal trisomy, both single and multiple, was the most common abnormality (207 cases, 63.5% of abnormalities). Trisomy 16 was the most frequent single trisomy, followed by trisomy 22, trisomy 15 and trisomy 21. The rate of chromosomal abnormalities of fetuses in early pregnancy (65.4%) was higher than that of fetuses in middle pregnancy (16.4%). There were 82 pregnant women with recurrent abortion, and the embryo with pathogenic CNVs was conceived in 62 (76%) cases, while the embryo with variants of unknown significance (VUS) in 12 (15%) cases. Among the 27 couples with a history of recurrent pregnancy loss (RPL), there were 4 (14.8%) couples with CNV abnormal in both partners, 16 (59.3%) couples with CNV abnormal only in the women, and 7 (25.9%) couples with CNV abnormal only in the men. These retrospective analyses of CNV-seq results provided a reference for genetic counseling of the relationship between VUS and RPL.

## INTRODUCTION

Spontaneous abortion (SA) is one of the most common pregnancy complications, with an incidence of 10% ∼ 15% in women of childbearing age. The incidence of recurrent spontaneous abortion (RSA) is 0.7% ∼ 1.9% ^12^. Chromosomal abnormalities are one of the important causes of spontaneous abortion, including chromosome number and structure abnormality. The incidence of chromosomal karyotype abnormalities in spontaneous abortion embryos is more than 50%, such as chromosome aneuploidy, polyploidy, mosaic monosomy/trisomy and so on ^3,4^. Recent studies have shown that chromosome copy number variation (CNV) may play a role in spontaneous abortion by influencing pregnancy-related genes or pathways^5^. Therefore, the importance of CNV in the cause of spontaneous abortion should be emphasized. The American College of Obstetricians and Gynecologists (ACOG), the Royal College of Obstetrics and Gynecology (RCOG), the American Society of Reproductive Medicine (ASRM) recommended that the chromosome of the abortions should be a routine clinical test, specially for RSA couples ^67^. This not only detect the cause of abortion, but also can identify whether husband and wife carry the underlying chromosome structural variation, and provide evidence for the choice of the method of reproduction.

Because of the low-throughput in the chromosome karyotype analysis, low-coverage massively parallel CNV sequencing (CNV-seq) has been rapidly applied in the field of spontaneous abortion. And the CNV-seq have many advantages such as wide detection range, precision, high throughput, low cost and simple operation ^8^. This study is expected to provide meaningful data for the genetic etiology of recurrent pregnancy loss. At the same time, it also provides some clinical basis for the genetic counseling in variants of uncertain significance CNV.

## Materials and methods

### Participants

There were 662 fetal samples of miscarriages and 54 peripheral blood sample collected from the Department of Obsterics and Gynecology, Zhongnan Hospital of Wuhan University, China, from 2018 to 2022. All parents consented to test voluntarily and provided signed informed consent. The gestational age at the time of miscarriage ranges from 4 to 29 weeks. The study was performed under the guidance of the Declaration of Helsinki and approved by the Ethics Committee of Zhongnan Hospital of Wuhan University (Ethics No.2023049K).

### CNV Sequencing and Data Analysis

CNV-seq was carried out by following the protocol mentioned in Xiya Zhou et al ^[9]^. Genomic DNAs were extracted using DNAeasy Kit (Qiagen, Valencia, CA, Unite States). All genomic samples for library construction were quantified using Qubit 3.0 (Invitrogen, Waltham, MA, USA). The qualified libraries were sequenced on the NextSeq 500 platform. Rawdata files were obtained from NextSeq 500, and then were demultiplexed and converted to fastq format using bcl2fastq software for downstream analysis. Adapters and reads with low quality were trimmed using fastp software. The BAM files were obtained by aligning the sequence reads to the reference (GRCh37/H19) with the use of the SpeedSeq. Additionally, duplicate reads were flagged in the BAM files to prevent downstream variant call errors, sample contamination and swaps using VerifyBamID. Circular binary segmentation (CBS) algorithm was used to remove low-quality base sequences carried out copy number analysis to find the reliability of chromosomal fragment variation according to Z-score and finally obtained the CNV situation on human 23 pairs of chromosomes.

### CNV classification principles

According to the joint consensus recommendation of ACMG and ClinGen, CNV is classified into five categories ^10^(pathogenic CNV, likely pathogenic CNV, variants of uncertain significance CNV, likely benign CNV, and benign CNV). Genomic variant databases including DGV (http://dgv.tcag.ca/dgv/app/homr), DECIPHER (http://decipher.anger.ac.uk), OMIM (http://www.omim.org), ClinGen (https://www.clinicalgenome.org/) and UCSC (http://genome.ucsc.edu/, hg19) were used as a reference source of CNV.

### Statistical Analysis

SPSS statistical software version 20.0 and GraphPad software version 19.0 were used for data analysis. Data was reported with the descriptive statistics method and measurement data was expressed as mean ± standard deviation (SD). Chi-square test was used to analyze the difference among the groups. A value of p < 0.05 was considered as statistically significant.

## Results

### Baseline characteristics and overall CNV results

A total of 662 aborted embryonic tissues were successfully analyzed by CNV-seq. The specimens tested include sporadic miscarriage (571 [86.2%]), recurrent pregnancy loss (82 [12.4%]) **(Table 1)**. The normal group included the samples with bCNV, lbCNV and undetected CNV, while the abnormal group includes the samples with pCNV and lpCNV. And the remained samples were grouped to VUS. Median maternal age was similar among those with normal or abnormal groups (30.2 years and 31.6 years, P> 0.01, respectively) **(Supplementary Figure 1)**. Fetal gestational age ranged from 4 weeks to 29 weeks (median of 11 weeks), the median gestational age for women conceived fetuses with pathogenic CNV was 9 weeks compared with 14 weeks in the normal group **(P<0**.**001, Supplementary Figure 1)**.

**Table 1.** Overall chromosomal results by clinical indication.

| Referral reason | bCNVs | pCNVs | lbCNVs | lpCNVs | VUS | Total, n(%) |
| --- | --- | --- | --- | --- | --- | --- |
| Miscarrige, n | 234 | 249 | 10 | 4 | 74 | 571(86.2) |
| RPL, n | 0 | 74 | 0 | 0 | 8 | 82(12.4) |
| Other, n | 5 | 3 | 0 | 0 | 1 | 9 (1.4) |
| Total, n (%) | 239 (36.1) | 326 (49.2) | 10 (1.5) | 4 (0.6) | 83 (12.6) | 662 |

bCNVs, benign CNVs; pCNVs, pathogenic CNVs; lbCNVs, likely benign CNVs; lpCNVs, likely pathogenic CNVs; VUS, variants of uncertain significance; RPL, recurrent pregnancy loss.

### Detailed classification of abnormal chromosomal findings

There were 326 (49.2%) samples with clinically significant pathogenic CNV, 83 (12.6%) samples with VUS CNV, 239 (36.1%) samples with benign CNV, 10 (1.5%) with likely benign CNV and 4 (0.6%) samples with likely pathogenic CNV **(Table 1)**. The abnormalities could also be classified to autosomal trisomy, autosomal monosomy, monosomy X, and segmental abnormalities **(Table 2 and Figure 1)**. Autosomal trisomy, both single and multiple, was the most common abnormality (207 cases, 63.5% of abnormalities) **(Table 2)**. These included 163 samples with a single trisomy (50.0%), 10 with multiple trisomies (3.07%) and 34 samples with single trisomy and segmental CNV (10.43%). Trisomy 16 was the most frequent single trisomy, followed by trisomy 22, trisomy 15 and trisomy 21 **(Figure 1)**. Autosomal monosomies were uncommon and accounted for 9 samples (2.76% of all abnormalities). Of these, three had isolated autosomal monosomy and six had additional abnormalities. Mosaicism was observed in two of the six cases (33.3%). Abnormalities involving chromosome X were common and complex. Among 45 cases (13.8% of abnormalities), isolated monosomy X was the most common and was observed in 40 cases (12.27%). Mosaicism was observed in 8 of the 40 cases (20%). In addition, polyploid was observed in 32 samples (9.82% of abnormalities); 30 cases were pure triploidy, in two triploid samples, a segmental abnormality was also observed. Segmental CNV were identified as the only abnormality in 33 cases (10.12%).

**Table 2.** Detailed description of abnormalities.

| Abnormality | Case, n | Frequency of abnormality among abnormal cases(%) (n=326) | Overall frequency (%) (n=662) | Notes |
| --- | --- | --- | --- | --- |
| Autosomal trisomy | 207 | 63.50 | 31.27 |  |
| Single | 163 | 50.00 | 24.62 | Mosaic in 9(5.5%) |
| Multiple | 10 | 3.07 | 1.51 |  |
| (Double trisomy) | (9) |  |  |  |
| (Three or more) | (1) |  |  |  |
| Autosoma trisomes + segmental | 34 | 10.43 | 5.14 | Mosaic in 5(14.7%) |
| Autosomal monosomy | 9 | 2.76 | 1.36 |  |
| Single | 3 | 0.92 | 0.45 |  |
| Autosomal monosomy + segmental CNV | 6 | 1.84 | 0.91 | Mosaic in 2(33.3%) |
| Monosomy X (Complete and partial) | 45 | 13.80 | 6.80 |  |
| Pure monosomy X | 40 | 12.27 | 6.04 | Mosaic in 8(20%) |
| Monosomy X + autosomal trisomy | 2 | 0.61 | 2.9 |  |
| Monosomy X + autosomal segmental CNV | 3 | 0.92 | 0.30 |  |
| Polyploidy | 32 | 9.82 | 4.83 |  |
| Triploidy | 30 | 9.20 | 4.53 |  |
| Triploidy + Segmental CNV | 2 | 0.61 | 0.30 |  |
| Segmental abnormalities (only) | 33 | 10.12 | 4.98 |  |

**Figure 1.**
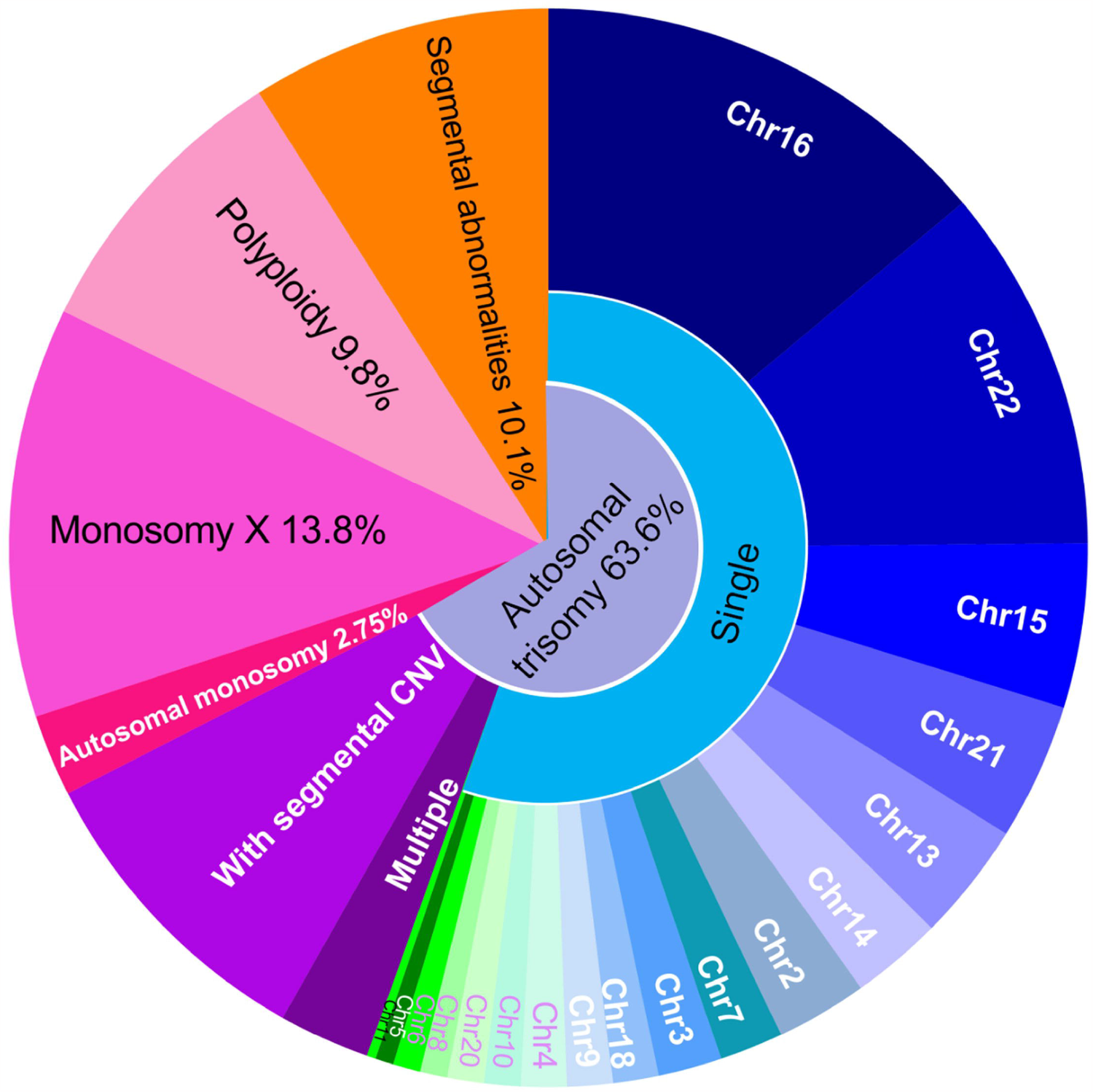
Distribution and relative frequencies of the major classes of abnormalities identified by CNV sequencing. Chr, chromosome; CNV, copy number variants;

The incidence of numerical chromosomal abnormalities in ≥ 35-year-old age pregnant women was significantly higher than < 35-year-old group (p=0.001, **Table 3**). Meanwhile, the rate of pCNV in ≥ 35-year-old age pregnant women increased significantly than < 35-year-old group (p<0.001, **Figure 2A**). The pCNV of abortion mainly detected in ≤ 12 weeks of gestation (95.4%), including ⩽ 6 weeks (11%), 7-8 weeks (36.7%), 9-10 weeks (29.7%) and 11-12 weeks (18%) **(Figure 2B)**. Compared with the normal group, the abortion carrying pCNV were rarely more than 13 weeks. On the other hand, the incidences of VUS CNV and normal group were not significantly different among gestational age groups **(Figure 2B)**.

**Table 3.** Comparison of CNVs results among different age pregnant women and gestational week of fetuses and amniotic fluid.

| CNVs results of fetuses | Average age of pregnant women |  |  |  | Gestational week |  |  |  |
| --- | --- | --- | --- | --- | --- | --- | --- | --- |
| | <35 years (n) | ≥35 years (n) | $\chi^2$ | *p* | Early pregnancy ( ≤12w ) (n) | Middle and late pregnancy( >12w ) (n) | $\chi^2$ | *p* |
| Normal | 206 (45.7%) | 33 (28.9%) | 10.781 | 0.0010 | 159 (34.6%) | 80 (83.6%) | 82.62 | $9.95 \times 10^{-20}$ |
| Chromosomal abnormalities | 245 (54.3%) | 81 (71.1%) |  |  | 310 (65.4%) | 16 (16.4%) |  |  |
| Chromosome number abnormalities | 188 (76.7%) | 68 (83.9%) | 1.971 | 0.160 | 258 (83.2%) | 11 (68.7%) | 1.90 | 0.168 |
| Chromosome structural abnormalities | 57 (23.3%) | 13 (16.1%) |  |  | 52 (16.8%) | 5 (31.3%) |  |  |
| CNVs results of amniotic fluid | <35 years (n) | ≥35 years (n) | $\chi^2$ | *p* | pregnancy ( ≤20w ) (n) | pregnancy ( >20w ) (n) | $\chi^2$ | *p* |
| Normal | 1107 (87.2%) | 423 (88.1%) | 0.295 | 0.587 | 720(83.8%) | 810(90.9%) | 20.185 | $0.7 \times 10^{-5}$ |
| Chromosomal abnormalities | 163 (12.8%) | 57 (11.9%) |  |  | 139(16.2%) | 81(9.1%) |  |  |
| Chromosome number abnormalities | 104 (63.8%) | 46 (80.7%) | 5.917 | 0.015 | 104(74.8%) | 46(56.8%) | 7.549 | 0.006 |
| Chromosome structural abnormalities | 59 (36.2%) | 11 (19.3%) |  |  | 35(25.2%) | 35(43.2%) |  |  |

**Figure 2.**
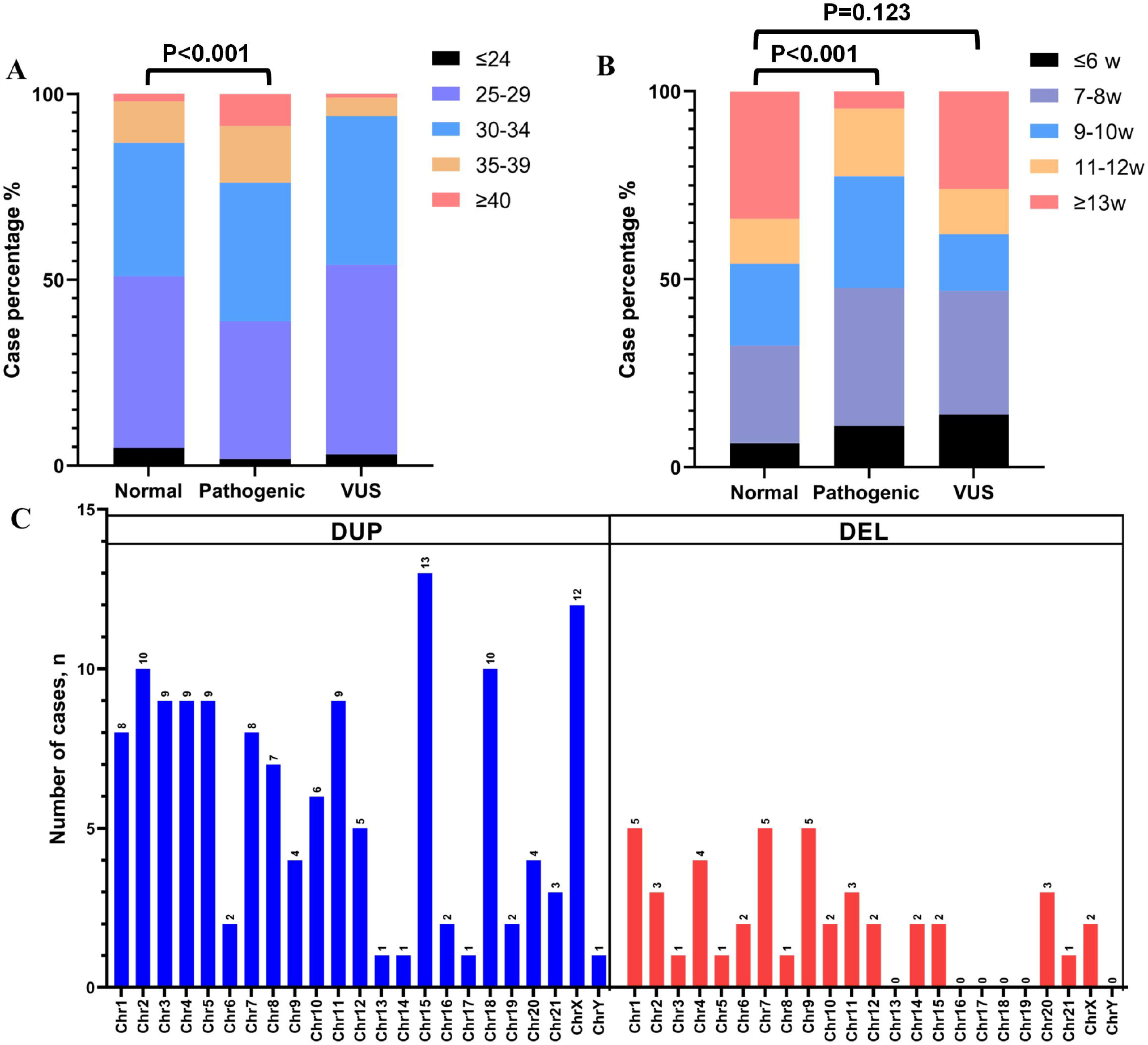
Chromosomal variation analysis. (A) Age distribution of numerical abnormalities of chromosomes in fetuses. (B) Gestational age distribution of numerical abnormalities of chromosomes in fetuses. (C) Distribution of duplication and deletion in VUS results. (D-E) Result of amniotic fluid CNV results in 1937 samples.

### Comparison of CNV Results Among Pregnant Women with Different Age and Gestation

In table 3, the incidence of numerical chromosomal abnormalities in ⩾35-year-old age pregnant women was significantly higher than <35-year-old group. However, the incidence of chromosome structural abnormalitie were not different between ⩾ 35 and < 35 year-old groups (p=0.325, Table 3). The rate of numerical chromosomal abnormalities were high (83.9%) in the⩾35 year-old age pregnant women, while the rate of structural chromosomal abnormalities was nominally lower (16.1%). The rate of chromosomal abnormalities of fetuses in early pregnancy (65.4%) was statistically higher than that of fetuses in middle pregnancy (16.4%) (p<0.001), whether it was chromosome structural abnormalitie or chromosome number abnormalities (Table 3). The classification of VUS included 75.5% duplication, 24.5% deletion. The most duplication was found on chromosome 15, followed by chromosome X and chromosome 18, and the most deletion was found on chromosome 1, 7 and 9. **(Supplementary Table 1, Figure 2C)**.

### Results of abortion tissue and peripheral blood CNV examination in couples with recurrent pregnancy loss

Among 82 recurrent abortions, there were 62 (76%) samples with pCNV, 12 (15%) samples with VUS and the remained samples without CNV abnormalities **(Figure 3A)**. Chromosome 16 and chromosome 22 were the most common chromosomal abnormalities, followed by triploid syndrome and segmental abnormalities **(Figure 3B)**. There were 27 couples which traced back to the source of CNV. All 27 couples had different levels of CNV **(Supplementary Table 2)**. About 3 of 27 (11.1%) couples carried pCNV/lpCNV. 22 of 27 (81.5%) couples carried VUS and the remaining couples carried benign CNV. Among the 27 couples, there were 4 (14.8%) couples with CNV abnormal in both partners, 16 (59.3%) couples with CNV abnormal only in the woman, and 7 (25.9%) couples with CNV abnormal only in the man. Of these, 68.4% was duplication, with the most occurrence in chromosomes 8 and X. 31.6% was deletion, with the most occurrence in chromosomes 6, 7 and 18. CNV-related genes were shown in **Supplementary Table 2**.

**Figure 3.**
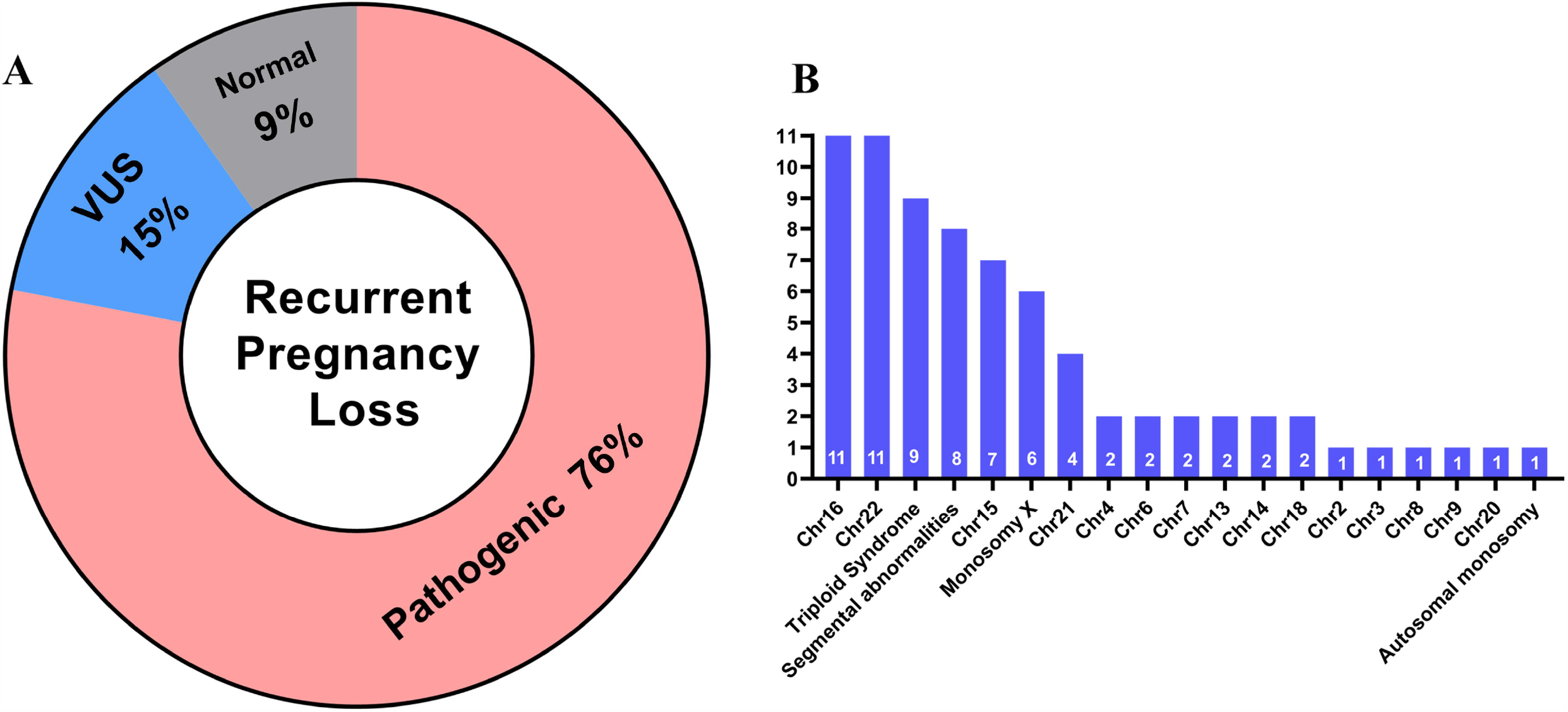
Recurrent pregnant loss and peripheral blood CNV analysis. (A-B) Results of fetuses in patients with recurrent pregnant loss. (C) Analysis of CNV results in people who came for genetic counseling with peripheral blood. (D) Analysis of peripheral blood CNV results in 54 couples with a history of abortion.

## Discussion

In this study, the CNV-seq results of 662 fetal specimens and 54 peripheral blood samples were retrospectively analyzed to provide reference for clinical consultation. The human genome of about 4.8%-9.5% populations contains CNV, more than 99% of which are benign variations or polymorphisms, but the other 1% of CNV can cause serious disease ^11^. Many fetuses with chromosomal microdeletion and microduplication syndrome have no ultrasound abnormalities in the first and second trimester, or only appear in the third trimester, which has missed the opportunity of prenatal diagnosis. Although NIPT-Plus technology has emerged in recent years, it has limited accuracy for CNV less than 5Mb. Anyway, further prenatal diagnosis is needed for verification. At present, as a widely used and mature technical means in clinical practice, chromosomal microarray analysis (CMA) and low-depth genome-wide copy number variation detection (CNV-Seq) can detect chromosomal microdeletions and microduplication.

Our study showed that the pathogenic CNV group was significantly lower than the normal group in the median gestational age. This result is similar to other studies ^12,13^. About 49.2% (326/662) aborted tissues carried clinically significant pathogenic CNV. 78% (64/82) cases with recurrent abortion were found to have pathogenic CNV. These results indicated that pathogenic CNV is one of the main causes of recurrent abortion. In our research, the trisomy variation mainly occurred on chromosomes 16 22 15 21 and 13, which were consistent with other studies ^14,15^. Alought chromosome 16 has more genes than chromosomes 13 18 21 22, trisomy 16 is the most common (about one third) autosomal trisomy found in abortuses. For the exact cause not well understood, chromosome 16 appears to be particularly vulnerable to nondisjunction. Meanwhile, trisomy 16 in abortuses shows little association with increasing maternal age^16^. On the other hand, numerical abnormalities were not detected on chromosomes 1 and 19 in our study. In the real world, Trisomy 1 and 19 syndrome is rare. This might be fatal for the embryonic development in Trisomy 1 and 19. Very few cases of trisomy 1 and 19 abortion have been reported ^17,18^.

In the comparison of chromosomal abnormalities between ⩾ 35 and < 35 year-old age pregnant women, we found that the rate of numerical chromosomal abnormalities was higher in the people ⩾ 35 year-old age. Because the probability of numerical chromosomal abnormalities increases with age, the rate of pCNV in ⩾ 35 old pregnant women increases significantly. Previous studies have shown that advanced age is an important factor causing chromosomal abnormalities in aborted embryos. It has been reported that oocytes of germ cells, with the increase of female age, have a longer stay in meiosis, which makes it more likely that lesions will occur. In addition, our research revealed that the abortion carrying pCNV were rarely more than 13 weeks. Most pCNV detected in ⩽ 12 weeks of gestation (95.4%). These results indicated that embryos carrying pCNV were aborted early in pregnancy, which was consistent with previous reports ^19^. Structural chromosomal abnormality is also one of main genetic factors in miscarriage, CNV in these chromosome contain many coding genes. If the CNV involves the increase or decrease of large gene fragments, and there are likely clinical manifestations of serious defects. In our study, pathogenic structural abnormalitie is more likely to lead early miscarriage, regardless of the age of the pregnant woman ^20,21^.

CNV-seq is a new generation of high-throughput sequencing technology that can simultaneously analyze hundreds of thousands to millions of DNA molecules at a time. It also provides coverage detection for the whole genome, chromosomal microdeletions and microduplications larger than 100kb can be detected . Although the resolution of CNV-seq technology is high, it also detected many CNV with unclear clinical significance. This brings challenges and difficulties to the interpretation of CNV in genetic counseling, specially for the spontaneous abortion ^22^. The Society of Obstetricians and Gynaecologists of Canada (SOGC)-Canadian College of Medical Geneticists (CCMG) recommended that variants of unknown significance (VUS) smaller than 500 Kb deletion or 1 Mb duplication not be routinely reported in the prenatal context ^23^. However, there are no professional guidelines for the reporting scope of CNV for miscarriages and recurrent pregnancy loss. In particular, for this population, whether VUS is the cause of recurrent abortion is often very difficult to consult. There was only a few literature analysis of the relationship between VUS and recurrent abortion ^13,24^. In the future, a large accumulation of CNV data will be needed to define which VUS cause miscarriages and recurrent pregnancy loss. These works will provided a reference for genetic counseling of assisted reproductive technology.

## Data Availability

All data produced in the present study are available upon reasonable request to the authors

## ETHICS STATEMENT

The study has been approved by the Ethics Committee of Zhongnan Hospital of Wuhan University, in accordance with the Declaration of Helsinki.

## AUTHOR CONTRIBUTIONS

ZF conceived the study. LJ and GN performed CNV tests. HH and YL performed all data analyses. ZF, HH, LJ wrote the manuscript; ZF, DJ, ZY and MJ executed the contents. GN, NY, QX revised the draft of the manuscript; JX, YL, SS, CY collected the clinical data. All authors reviewed and approved the final manuscript.

## Figure Legend

**Supplementary Figure 1.** Distribution of CNV results compared with median pregnancy gestational age and median maternal age. Gestational age (GA) information was available for analysis for 662 products of conception samples.

