## Supplementary Figure 1 V2 for "Chromosomal copy number variations in miscarriages and the geneic counseling of recurrent pregnancy loss"

| Case  Supplementary Table 1 Characteristics of VUS in CNV detection of abortion samples (Deletion or Duplication size >500kbp) | Chromosome | Dup/Del | Size (kbp) | Location | Isolated VUS or accompanied by a pathogenic mutation | Morbid Genes |
| --- | --- | --- | --- | --- | --- | --- |
| 1 | 1 | Del | 7600 | 1p36.33(820001-8420000) | Isolated VUS | ISG15,AGRN,DVL1,VWA1,TNFRSF4,B3GALT6,ATAD3A,GNB1,GABRD,SKI,PEX10,PANK4,PRDM16,TP73,SMIM1,CEP104,NPHP4,CHD5,ESPN,PLEKHG5,CAMTA1,PER3,PARK7 |
| 1 | 15 | Del | 900 | 15q15.1(40980001-41880000) | Isolated VUS | CHP1,NDUFAF1,MAPKBP1 |
| 2 | 3 | Dup | 1000 | 3p21.31(46060001-47060000) | Isolated VUS | CCR2,CCR5,TDGF1,TMIE,MYL3,PTH1R,NBEAL2,SETD2 |
| 3 | 15 | Dup | 1860 | 15q22.2(59560001-60380000) | Isolated VUS | BNIP2,FOXB1 |
| ~~4~~ | ~~15~~ | ~~Dup~~ | ~~780~~ | ~~15q11.2(22740001-23520000)~~ | ~~Isolated VUS~~ | ~~NIPA1~~ |
| 5 | 1 | Dup | 800 | 1p36.33(1060000-1800000) | Isolated VUS | TNFRSF4,B3GALT6,DVL1,VWA1,ATAD3A,TMEM240 |
| 6 | 11 | Dup | 740 | 11p15.3-2(12120001-12860000) | Isolated VUS | TEAD1 |
| 7 | 18 | Dup | 580 | 18p11.23(7160001-7740000) | Isolated VUS | PTPRM |
| 8 | 18 | Dup | 520 | 18p11.31-3(7080001-7600000) | Isolated VUS | PTPRM |
| 9 | 22 | Dup | 2300 | 22q11.21(18880001-21180000) | Isolated VUS | PRODH,SLC25A1,CDC45,GP1BB,TBX1,TXNRD2,TANGO2,COMT,RTN4R,SCARF2,P14KA,SERPIND1,SNAP29,LZTR1 |
| 10 | 15 | Dup | 540 | 15q13.3(31900001-32440000) | Isolated VUS |  |
| 11 | 5 | Dup | 1020 | 5q21.3-22.1(109540001-110560000) | Isolated VUS | MAN2A1 |
| 12 | 2 | Dup | 800 | 2p24.3(13540000-14340000) | Isolated VUS |  |
| 13 | 1 | Del | 1340 | 1q41(222680000-224020000) | Isolated VUS | TLR5 |
| 14 | 21 | Dup | 620 | 21q22.11(34560000-35180000) | Isolated VUS | RUNX1 |
| 15 | 9 | Dup | 2280 | 9p21.3-2(24340000-26620000) | Isolated VUS |  |
| 16 | 10 | Del | 1900 | 10q26.3(133540000-135440000) | Isolated VUS | EBF3,NKX6-2,TUBGCP2,ECHS1,SYCE1 |
| 17 | 12 | Dup | 880 | 12q21.33(90920000-91800000) | Isolated VUS | KERA,DCN |
| 18 | 14 | Dup | 820 | 14q32.33(105640000-106460000) | Isolated VUS | IGHG2,IGHM |
| 19 | 20 | Dup | 2720 | 20p13(60000-2780000) | Isolated VUS | RBCK1,TBC1D20,CSNK2A1,SLC52A3,RSPO4,PDYN,TGM3,TGM6,SNRPB,NOP56,IDH3B |
| 20 | 20 | Dup | 920 | 20p13(2780000-3700000) | Isolated VUS | VPS16,AVP,DDRGK1,ITPA,SLC4A11 |
| 21 | 7 | Del | 1920 | 7q36.1(150620001-152540000) | Isolated VUS | GIMAPS,KCNH2,CDK5,NOS3,PRKAG2,KMT2C |
| 21 | 15 | Dup | 900 | 15q13.3(32020001-32920000) | Isolated VUS | GREM1 |
| 22 | 4 | Dup | 1060 | 3q32.3(168320001-169380000) | Isolated VUS | MECOM |
| 23 | 2 | Dup | 700 | 2p25.3(1100001-1800000) | Isolated VUS | TPO,PXDN,MYT1L |
| 24 | 3 | Dup | 500 | 3p12.3(78400001-78900000) | Isolated VUS | ROBO1 |
| 25 | X | Dup | 800 | Xp11.23-22(49080001-49880000) | Isolated VUS | WDR45,PRICKLE3,CCDC22,SYP,FOXP3,CACNA1F |
| 26 | 12 | Dup | 980 | 12q21.31(80660001-81640000) | Isolated VUS | MYF5 |
| 27 | 18 | Dup | 700 | 19p11.32(87380001-87680000) | Isolated VUS |  |
| 28 | 15 | Dup | 540 | 15q26.3(101860001-102400000) | Isolated VUS | IGF1R |
| 29 | 8 | Dup | 860 | 8q11.1(46880000-47740000) | Isolated VUS | SPIDR |
| 30 | 8 | Dup | 860 | 8q11.1(46880000-47740000) | Isolated VUS | SPIDR |
| 31 | 22 | Dup | 1120 | 22q11.23(23880000-25000000) | Isolated VUS | MIF,SPECC1L,UPB1,GGT1 |
| 32 | 2 | Dup | 1860 | 2p16.2-1(53780000-55640000) | Isolated VUS | SPTBN1,CCDC88A |
| 33 | 5 | Dup | 1100 | 5q21.3-1(108840000-109940000) | Isolated VUS | MAN2A1 |
| 34 | X | Dup | 1100 | Xq11.1-2(62920000-64020000) | Isolated VUS | ARHGEF9 |
| 35 | 2 | Dup | 760 | 2p24.3(13580001-14340000) | Isolated VUS |  |
| 35 | 15 | Dup | 800 | 15q11.2(22300001-23100000) | Isolated VUS | NIPA1 |
| 36 | 21 | Del | 10820 | 21q11.2-q21.2(15060001-25880000) | VUS accompanied by a pathogenic mutation | TMPRSS15,NIRIP1 |
| 37 | 18 | Dup | 1680 | 18p11.21(11640001-13320000) | VUS accompanied by a pathogenic segmental | GNAL,TUBB6,AFG3L2,PSMG2 |
| 38 | 5 | Dup | 980 | 5p14.3(20400001-21380000) | VUS accompanied by a autosomal trisomy |  |
| 39 | 2 | Del | 1720 | 2q13(111380001-113100000) | VUS accompanied by a autosomal trisomy | ANAPC1,MERTK,POLR1B,CKAP2L,IL1B,IL37,IL36RN,IL1RN |
| 40 | 8 | Dup | 1800 | 8p11.21(39880000-41680000) | VUS accompanied by a pathogenic segmental | SFRP1,ANK1 |
| 41 | X | Dup | 640 | Xp22.2(14140001-14780000) | VUS accompanied by a autosomal trisomy | GLRA2,FANCB |
| 42 | 5 | Dup | 720 | 5p14.3(20400001-21120000) | VUS accompanied by a autosomal trisomy |  |
| 43 | 8 | Del | 1440 | 8p12(31280001-32720000) | VUS accompanied by a pathogenic segmental | NRG1 |
| 44 | 4 | Dup | 740 | 4p15.1(33260001-34860000) | VUS accompanied by a pathogenic segmental |  |
| 44 | 4 | Del | 3520 | 4q35.2(187360001-190880000) | VUS accompanied by a pathogenic segmental |  |
| 44 | 4 | Dup | 1600 | 4p15.1(32520001-3260000) | VUS accompanied by a pathogenic segmental |  |
| 45 | 15 | Dup | 560 | 15q13.3(31900001-32460000) | VUS accompanied by a autosomal trisomy |  |
| 46 | 4 | Dup | 1120 | 4q12-13.1(58600001-59720000) | VUS accompanied by a autosomal trisomy |  |
| 46 | 8 | Dup | 640 | 8q13.2(68400001-69040000) | VUS accompanied by a autosomal trisomy |  |
| 47 | 12 | Dup | 560 | 12p12.1(23340000-23900000) | VUS accompanied by a autosomal trisomy | SOX5 |
| 48 | 3 | Dup | 620 | 3p13-12.3(73880000-74500000) | VUS accompanied by a autosomal trisomy |  |
| 49 | 11 | Dup | 1240 | 11q21-q22.1(96800000-98040000) | VUS accompanied by a autosomal trisomy |  |
| 50 | 7 | Del | 1780 | 7q36.3(157360000-159138663) | VUS accompanied by a pathogenic segmental | DNAJB6,NCAPG2,DYNC2I1 |
| 51 | 8 | Dup | 500 | 8q22.3(103280000-103780000) | VUS accompanied by a pathogenic segmental | FZD6,CTHRC1,SLC25A32,RIMS2 |
| 52 | 10 | Del | 2040 | 10p12.31-p12.2(21540000-23580000) | VUS accompanied by a autosomal trisomy | MLLT10,PTF1A |
| 53 | 2 | Dup | 960 | 2p22.3(34040000-35000000) | VUS accompanied by a pathogenic segmental |  |
| 54 | X | Dup | 580 | Xq23-24(116260000-116840000) | VUS accompanied by a autosomal trisomy |  |
| 55 | 4 | Dup | 800 | 4q31.3(151720000-152520000) | VUS accompanied by a autosomal trisomy | GATB,FBXW7 |
| 56 | 19 | Dup | 620 | 19q11-12(28020000-28640000) | VUS accompanied by a autosomal trisomy |  |
| 57 | 3 | Dup | 1260 | 3q28(190380000-191640000) | VUS accompanied by a autosomal trisomy | CLDN16,CCDC50 |
| 58 | Y | Dup | 600 | Yp11.2(9380000-9980000) | VUS accompanied by a pathogenic segmental |  |
| 59 | 3 | Dup | 580 | 3q13.33(120780000-121360000) | VUS accompanied by a autosomal trisomy |  |
| 59 | 15 | Dup | 560 | 15q13.3(31880000-32440000) | VUS accompanied by a autosomal trisomy |  |
