## Supplementary Table 2 V2 for "Chromosomal copy number variations in miscarriages and the geneic counseling of recurrent pregnancy loss"

| Case | Parental origin of CNV  Supplementary Table 2 Characteristics of peripheral blood CNVs in the RPL | Chr | Dup/Del | Size (kbp) | Location | CNV result | Adverse pregnancy history | Genes in region/gaps | Description of test results |
| --- | --- | --- | --- | --- | --- | --- | --- | --- | --- |
| 1 | Maternal | 18 | Del | 4740 | 18q21.33q22.1(61540001-66280000) | lpCNV | Miscarriage 3 times | PIGN,TNFRSF11A,BCL2,KDSR,SERPINB7,SERPINBB | The deletion of the 4.74Mb region at q21.33-q22.1 of chromosome 18 has been reported in the literature. 46,XX,t(1;2;18)(q32;Q24.2;q22.1).ish del(18)(q22.1q22.1) (RP11-526H11x1).arr [hg38] 18q22.1(65,612,478 -- 68,326,910)×1. The main clinical manifestations of the patient were mild facial features,Mild learning difficulties and mild mental retardation [PMID:26681178]. |
| 1 | Paternal | 22 | Dup | 300 | 22q11.22(22320000-22620000) | Benign | Miscarriage 3 times |  | This segment is polymorphic |
| 2 | Maternal | 13.14.21 | Dup | 150-600 | 13q31.1（82670001-83270000）14q24.2（70400001-70550000）21q21.1（18561194-19061193） | VUS | Miscarriage 3 times | SLC8A3,C21orf37,BTG3,CXADR | No clear pathogenic information and literature reports related to these segments were found. |
| 2 | Paternal | 2.8.11 | Del | 150-600 | 11q21（96617001-96767000）  2q24.1（156579852-157179851）8q21.13（82465101-82865100） | VUS | Miscarriage 3 times | SLC10A5,IMPA1,SNX16,CHMP4C,ZFAND1 | No clear pathogenic information and literature reports related to these segments were found. |
| 3 | Maternal | 7 | Del | 360 | 7q11.21(64460001-64920000) | Benign | Miscarriage 3 times |  | This segment is polymorphic |
| 3 | Paternal | / | / | / | / | / | Miscarriage 3 times |  |  |
| 4 | Maternal | 7 | Dup | 400 | 7q31.1(110020000-110420000) | VUS | Miscarriage 2 times and birth a deformed fetus once | IMMP2L | No clear pathogenic information and literature reports related to this segment were found.DGV database queried the record of 1 normal person carrying this repeat fragment in the 17421 sample group. |
| 4 | Paternal | 6/9 | Dup | 220/260 | 6q21(113400000-113620000)  9q21.32-33(86820001-8780000) | VUS | Miscarriage 2 times and birth a deformed fetus once | NTRK2 | The duplicated 0.26Mb region at q21.32-q21.33 of  chromosome 9 was not found in the definitive pathogenic information and literature reports related to this segment. DGV database queried the record of 1 normal person carrying this repeat fragment in 29084 samples. |
| 5 | Maternal | / | / | / | / | / | Miscarriage 2 times and birth a deformed fetus(missing limbs) once |  |  |
| 5 | Paternal | 8 | Dup | 260 | 8q13.3(71260000-71520000) | VUS | Miscarriage 2 times and birth a deformed fetus(missing limbs) once | EYA1,TRAPPC2P2 | There is no report on this segment |
| 6 | Maternal | / | / | / | / | / | Miscarriage 3 times |  |  |
| 6 | Paternal | 8 | Dup | 340 | 8q24.22(131760001-132100000) | VUS | Miscarriage 3 times | EFR3A | DGV database queried the record of 1 normal person carrying this repeat fragment in 29084 samples. |
| 7 | Maternal | 9 | Dup | 1580 | 9q31.1(104520001-106100000) | VUS | Miscarriage 2 times and hydatidiform mole once | ABCA1,SLC44A1,FKTN,TAL2,TMEM38B | There is no report on this segment |
| 7 | Paternal | / | / | / | / | / | Miscarriage 2 times and hydatidiform mole once |  |  |
| 8 | Maternal | / | / | / | / | / | Miscarriage 3 times |  |  |
| 8 | Paternal | Y/15 | Dup | 700/520 | Yp11.2(7420001-8120000)  15q21.3(54820001-55340000) | VUS | Miscarriage 3 times | RAB27A,PIGB | There is no report on this segment |
| 9 | Maternal | / | / | / | / | / | Miscarriage 2 times and neonates CNV abnormal |  |  |
| 9 | Paternal | 14 | Del | 460 | 14q11.2(22540001-23000000) | Benign | Miscarriage 2 times and neonates CNV abnormal |  |  |
| 10 | Maternal | / | / | / | / | / | Miscarriage 2 times and birth a deformed fetus(Cleft lip and palate) once |  |  |
| 10 | Paternal | 12 | Del | 1080 | 12p11.21(33100001-34180000) | VUS | Miscarriage 2 times and birth a deformed fetus(Cleft lip and palate) once | ALG10,SYT10 | Similar DECIPHER Patient:407549, phenotype: abnormality of the face;autistic behavior; intellectually disability (moderate) |
| 11 | Maternal | X | Dup | 220 | Xq26.3(134760001-134980000) | VUS | Miscarriage 3 times |  | There is no report on this segment |
| 11 | Paternal | / | / | / | / | / | Miscarriage 3 times |  |  |
| 12 | Maternal | X | Dup | 120 | Xp11.4(42140001-42260000) | VUS | Miscarriage 3 times |  | There is no report on this segment |
| 12 | Paternal | / | / | / | / | / | Miscarriage 3 times |  |  |
| 13 | Maternal | 7 | Dup | 540 | 7q36.3(158400001-158940000) | VUS | Miscarriage 3 times | NCAPG2,DYNC2I1,ESYT2,PTPRN2 | There is no report on this segment |
| 13 | Paternal | / | / | / | / | / | Miscarriage 3 times |  |  |
| 14 | Maternal | / | / | / | / | / | / | / | / |
| 14 | Paternal | X | Dup | 1700 | Xp22.31（6460001-8160000） | VUS | Miscarriage 3 times | HDHD1,STS,VCX,PNPLA4,VCX2 | ClinGen database showed this region as benign variation (TS=40:Dosage sensitivity unlikely). But the repeat also contains a gene that codes for the VCX2 protein. |
| 15 | Maternal | X/5/6 | Dup | 380 | Xq12(67240001-67780000)  5q14.1(78340001-78980000)  6q14.1(80100000-80420000) | VUS | Miscarriage 2 times and birth a deformed fetus once | AR,ARSB,BCKDHB | There is no report on this segment |
| 15 | Paternal | / | / | / | / | / | Miscarriage 2 times and birth a deformed fetus once |  |  |
| 16 | Maternal | / | / | / | / | / | Miscarriage 3 times |  |  |
| 16 | Paternal | 12 | Dup | 120 | 12q13.12(49380001-49500000) | VUS | Miscarriage 3 times | SPATS2 | There is no report on this segment |
| 17 | Maternal | 1 | Dup | 120 | 1p31.1(73220001-73340000) | VUS | Miscarriage 3 times |  | Two records of normal people carrying this repeat fragment were found in DGV database, and the carrying ratio was 1/29084 |
| 17 | Paternal | / | / | / | / | / | Miscarriage 3 times |  |  |
| 18 | Maternal | 15/17 | Del/Dup | 340/380 | 15q11.2(22760000-23100000)  17q12(35440000-35820000) | pCNV/VUS | Miscarriage 2 times and birth a deformed fetus once | NIPA1,SLFN14,PEX12,GAS2L2,TAF15 | This segment contains the entire 15q11.2 recurrent region (BP1-BP2)(includes NIPA1)[ISCA ID:ISCA-37448],There is ample evidence (Haploinsufficiency score:3) shows that the single time of insufficiency in this region is related to the neural developmental insufficiency (2A, score 1) |
| 18 | Paternal | / | / | / | / | / | Miscarriage 2 times and birth a deformed fetus once |  |  |
| 19 | Maternal | 16 | Dup | 1180 | 16p13.11(15120000-16300000) | lpCNV | Miscarriage 3 times | NDE1,MYH11,ABCC1,ABCC6 | This recurrent region contains all of the 16p13.11 recurrent region (BP2-BP3) (includes MYH11)[ISCA ID:ISCA-37415], which is a recurrent region susceptible to neurocognitive disorders (autism, intellectual impairment, multiple congenital malformation).Partial evidence (Triplosensitivity score:2) supports triplosensitivity involvement in this region |
| 19 | Paternal | / | / | / | / | / | Miscarriage 3 times |  |  |
| 20 | Maternal | 10 | Del | 220 | 10q21.1(59560000-59780000) | VUS | Miscarriage 3 times(trisomy 6, trisomy 22, trisomy 16) |  | DGV and gnomAD databases queried several records of natural population carrying fragments of this missing region, the proportions were 5/771, 2/369, 3/3017, etc. (section4, score 0). |
| 20 | Paternal | / | / | / | / | / | Miscarriage 3 times(trisomy 6, trisomy 22, trisomy 16) |  |  |
| 21 | Maternal | 18 | Del | 260 | 18p11.23(7900000-8160000) | VUS | Miscarriage 3 times | PTPRM | There is no report on this segment |
| 21 | Paternal | 10 | Dup | 180 | 10q21.1(56520000-56700000) | VUS | Miscarriage 3 times |  | There is no report on this segment |
| 22 | Maternal | 3/8 | Del/Dup | 240/600 | 3p22.1(42740000-42980000)  8p23.3(480000-1080000) | VUS | Miscarriage 3 times and biochemical pregnancy once | TDRP,ERICH1,DLGAP2 | There is no report on this segment |
| 22 | Paternal | / | / | / | / | / | Miscarriage 3 times and biochemical pregnancy once |  |  |
| 23 | Maternal | 4/5/16 | Dup | 560/400/260 | 4p16.3(3300001-3860000)  5q23.1(115260001-115660000)  16p12.3(19960001-20220000) | VUS | Miscarriage 2 times and biochemical pregnancy once | DOK7,LRPAP1,ADRA2C | Repeat the 0.26Mb region at p12.3 on chromosome 16;DGV database queried the record of 1 normal person carrying this repeat fragment in 2 sample groups. |
| 23 | Paternal | / | / | / | / | / | Miscarriage 2 times and biochemical pregnancy once |  |  |
| 24 | Maternal | 2 | Dup | 460 | 2q32.3(19564000-19610000) | VUS | Miscarriage 2 times and biochemical pregnancy once |  | There is no report on this segment |
| 24 | Paternal | / | / | / | / | / | Miscarriage 2 times and biochemical pregnancy once |  |  |
| 25 | Maternal | 6 | Del | 180 | 6q16.3(101220001-101400000) | VUS | Miscarriage 3 times | GRIK2 | There is no report on this segment |
| 25 | Paternal | / | / | / | / | / | Miscarriage 3 times |  |  |
| 26 | Maternal | 6 | Del | 120 | 6q22.31(124200001-124320000) | VUS | Miscarriage 5 times |  | There is no report on this segment |
| 26 | Paternal | / | / | / | / | / | Miscarriage 5 times |  |  |
| 27 | Maternal | 7 | Dup | 620 | 7q33(133500000_134120000) | VUS | Miscarriage 4 times | :LRGUK,EXOC4 | There is no report on this segment |
| 27 | Paternal | / | / | / | / | / | Miscarriage 4 times |  |  |
